# Comparative Risk of Suicide-Related Events Across Antidepressant Classes in Autistic Individuals: A New-User Active Comparator Cohort Study

**DOI:** 10.64898/2026.10.21.26366126

**Authors:** Chen Jiang, Ruihan Huang, Safaa Eldeeb, Hui Wang, Zheng Chole Zeng, Kelly B Beck, Neal D Ryan, Carla A Mazefsky, LiRong Wang

**Affiliations:** Department of Pharmaceutical Sciences, University of Pittsburgh School of Pharmacy, Pittsburgh, PA, USA; Department of Biostatistics and Health Data Science, University of Pittsburgh School of Public Health, Pittsburgh, PA, USA; Department of Psychiatry, University of Pittsburgh, Pittsburgh, Pennsylvania, USA; Lumbrita LLC, Los Gatos, CA, USA

**Keywords:** Autism Spectrum Disorder, Antidepressive Agents, Suicide, Pharmacoepidemiology, Cohort Studies, Electronic Health Records

## Abstract

**Introduction:** Comparative evidence on suicide-related outcomes across antidepressant treatments among autistic individuals is limited. We examined associations between antidepressant treatment class and suicide-related events (SREs) among individuals on the autism spectrum initiating antidepressants.

**Methods:** We conducted a retrospective new-user, active-comparator cohort study using the nationwide Truveta database of electronic health records. Autistic individuals initiating antidepressants after an autism spectrum diagnosis were classified by initial treatment as a selective serotonin reuptake inhibitor (SSRI), atypical antidepressant, serotonin-norepinephrine reuptake inhibitor (SNRI), or tricyclic antidepressant (TCA). Pairwise propensity score overlap weighting was used to balance baseline characteristics, and associations with SREs during as-treated follow-up were estimated using weighted Cox proportional hazards models.

**Results:** Among 42,441 autistic antidepressant initiators, atypical antidepressants (hazard ratio [HR], 0.787; 95% confidence interval [CI], 0.632–0.979) and TCAs (HR, 0.487; 95% CI, 0.263–0.902) were associated with lower observed SRE hazards compared with SSRIs, whereas no clear difference was observed for SNRIs (HR, 0.948; 95% CI, 0.651–1.381). In agent-specific analyses, trazodone was associated with a lower observed SRE hazard compared with SSRIs, whereas no clear differences were observed for bupropion or mirtazapine. Exploratory age-stratified analyses suggested a lower hazard for atypical antidepressants versus SSRIs among individuals younger than 18 years, although evidence of effect modification was not consistent across alternative specifications of age.

**Discussion:** Among autistic individuals initiating antidepressants, observed SRE hazards differed across some antidepressant treatment comparisons. The class-level association for atypical antidepressants was not observed consistently across its individual agents, and age-related differences warrant further investigation.

**Lay Summary:** Autistic people are often treated with antidepressants, but little is known about whether different types of antidepressants are associated with differences in rates of suicide-related events. In this large real-world study, we compared several commonly used antidepressant treatments among autistic individuals and found that some treatment groups had lower observed rates of suicide-related events than others, although these findings should be interpreted cautiously because treatment choice may reflect differences between patients that cannot be fully measured in health records. The findings may help inform future research on the safety of antidepressant treatment in autistic people.

## 1. Introduction

Prevalence of co-occurring mental health conditions among autistic people is considerably higher than non-autistic people. (Hodges et al., 2020). Depression is particularly common among autistic people, especially during adolescence and adulthood. (Hudson et al., 2019; Pezzimenti et al., 2019). Meta-analytic estimates place the pooled prevalence of depression at 11-15% (Hudson et al., 2019; Lai et al., 2019), although rates differ markedly by assessment approach, increasing to 28% lifetime prevalence when assessed via clinical interview and 49% lifetime prevalence when based on self-report measures (Hudson et al., 2019). Autistic people are also at elevated risk for suicide-related outcomes (Brown et al., 2024). Specifically, death by suicide is three times more common in autistic people than non-autistic people (Santomauro et al., 2024). Autistic individuals also experience substantially elevated rates of suicidal ideation, suicidal behavior, and self-harm-related emergency care compared with the general population (Huntjens et al., 2024; Lai et al., 2023).

Given the high prevalence of depression and suicide-related outcomes in autistic people, anti-depressant medications are commonly prescribed. However, prescribing patterns of antidepressant treatment among autistic people are heterogeneous and include selective serotonin reuptake inhibitors (SSRIs), atypical antidepressants, serotonin-norepinephrine reuptake inhibitors (SNRIs), and tricyclic antidepressants (TCAs). Evidence from randomized antidepressant trials in autistic people remains sparse, and interpretation is hindered by methodological weaknesses and mixed findings across studies, precluding definitive conclusions about treatment effectiveness and adverse effects (Deb et al., 2021).

The potential relationship between antidepressant treatment and suicide-related events (SREs) has been extensively investigated in the general population. (Kozhevnikova et al., 2026; Reeves & Ladner, 2010). Safety concerns have been particularly prominent in younger populations, leading to an FDA boxed warning regarding suicidal thoughts and behaviors associated with antidepressant treatment (Stone et al., 2009). An umbrella review found evidence of an association between antidepressant use and suicide attempt or completion among children and adolescents, although this evidence was attenuated after accounting for confounding by indication (Dragioti et al., 2019). Previous studies have also reported variation in suicide-related outcomes across age groups, treatment indications, and individual antidepressant agents, with inconsistent findings across observational studies (Coupland et al., 2015; Pompili et al., 2010; Schetz et al., 2024; Schneeweiss et al., 2010; Wu et al., 2017). A recent population-based study examined the association between antidepressant initiation and suicide mortality among individuals with ASD but did not compare different antidepressant treatments (Chen et al., 2026).

Despite the high prevalence of co-occurring mental health conditions, frequent antidepressant use, and elevated underlying risk of suicide-related outcomes among autistic people, autism-specific evidence regarding suicide-related outcomes across antidepressant treatment remains limited (Manter et al., 2025). To address this knowledge gap, we conducted a new-user, active-comparator cohort study to compare the risk of SREs among autistic people initiating different classes of antidepressants. An active-comparator design was used to compare alternative antidepressant treatments among treated individuals and to reduce confounding that may arise when comparing antidepressant-treated and untreated individuals (Lund et al., 2015). We evaluated pairwise comparisons among SSRIs, SNRIs, atypical antidepressants, and TCAs and additionally examined the robustness of the findings across alternative study specifications and whether associations varied by age or sex.

## 2. Methods

### 2.1 Data Source and Study Design

This retrospective new-user, active-comparator cohort study used longitudinal, deidentified EHR data available from Truveta, a large-scale US healthcare database. In the resulting analytic cohort, eligible antidepressant treatment initiation dates ranged from March 31, 2008, through February 28, 2026, and follow-up was administratively censored on March 6, 2026. Truveta includes data from more than 130 million patients receiving care across more than 900 hospitals and 20,000 clinics in the United States and provides structured information on demographics, diagnoses, encounters, medication dispensing, and mortality (Burkhardt et al., 2026). The study compared the risk of documented suicide-related events among autistic individuals who initiated different antidepressant classes and were followed using an as-treated strategy.

### 2.2 Study Population

The study population included individuals of all ages, including children and adults, with at least one documented diagnosis of autism spectrum disorder (ASD) before antidepressant treatment initiation. Autism spectrum diagnoses were identified using qualifying diagnosis codes from the International Classification of Diseases, Tenth Revision, Clinical Modification (ICD-10-CM), International Classification of Diseases, Ninth Revision, Clinical Modification (ICD-9-CM), or Systematized Nomenclature of Medicine-Clinical Terms (SNOMED CT). The autism spectrum index date was defined as the start date of the earliest encounter linked to a qualifying autism spectrum condition.

Eligible individuals were required to have an antidepressant dispensing record on or after the autism spectrum diagnosis index date and sufficient observable healthcare history before treatment initiation. The treatment index date was defined as the first eligible antidepressant dispensing date on or after the autism spectrum diagnosis date, provided that no antidepressant dispensing was observed during the preceding 180 days. Individuals were required to initiate exactly one antidepressant class on the index date and to have a valid, positive days-supply value for the index dispensing. Individuals with a documented SRE or recorded death on or before the treatment index date were excluded to ensure valid at-risk follow-up for the first post-index SRE.

### 2.3 Antidepressant Exposure

Antidepressant exposure was defined using pharmacy dispensing records from Truveta. The initial treatment class was determined from all qualifying antidepressant dispensing on the treatment index date. The source cohort included selective serotonin reuptake inhibitors (SSRIs), serotonin-norepinephrine reuptake inhibitors (SNRIs), tricyclic antidepressants (TCAs), monoamine oxidase inhibitors (MAOIs), atypical antidepressants (bupropion, mirtazapine, and trazodone), and serotonin modulators. Comparative analyses were restricted to SSRIs, atypical antidepressants, SNRIs, and TCAs because the MAOI and serotonin-modulator groups were too small for stable pairwise estimation.

For each dispensing of the index antidepressant class, the exposure period began on the dispensing date and ended on the dispensing date plus the recorded days-supply minus 1 day. Overlapping records were consolidated, and consecutive records separated by no more than 30 days were combined into one continuous exposure episode. The end of the episode included the 30-day grace period. Initiation of an antidepressant from a class different from the index treatment class was considered a treatment change and resulted in censoring.

### 2.4 Suicide-Related Outcomes

The primary outcome was the first documented SRE after the treatment index date, identified using prespecified ICD-9-CM, ICD-10-CM, and SNOMED CT concepts encompassing suicidal ideation, suicide attempt, intentional self-harm, and intentional self-poisoning. The ICD-10-CM coding framework was consistent with established surveillance definitions for suicide attempt and intentional self-harm (Hedegaard et al., 2018).

Follow-up began on the treatment index date and continued until the first suicide-related event or censoring. Censoring occurred at the earliest of the following: initiation of an antidepressant from a class different from the index treatment class, indicating a treatment change; the end of the index-treatment exposure episode, including the 30-day grace period, indicating treatment discontinuation; death; 2 years after the treatment index date; or the end of the available study data. A suicide-related event was counted only when it occurred during this follow-up period.

### 2.5 Covariates

Baseline covariates were selected a priori based on prior literature and clinical relevance as potential predictors of antidepressant treatment selection and suicide-related outcomes (Brown et al., 2024; Jobski et al., 2017; Kim et al., 2024). Demographic covariates included age at the treatment index date, sex, and race. Age at index was calculated from the recorded date of birth and treatment index date. Co-occurring conditions included depression, attention-deficit/hyperactivity disorder, substance use disorder, schizophrenia spectrum disorders, bipolar disorder, anxiety, obsessive-compulsive disorder, posttraumatic stress disorder, eating disorders, personality disorders, intellectual disability, oppositional defiant or conduct disorder, intermittent explosive disorder, epilepsy, chronic pain, sleep-wake disorders, obesity or overweight, vision impairment, hearing impairment, and other sensory impairments. A co-occurring condition was considered present if a qualifying record was documented during the 365-day baseline period.

Baseline medication use was also assessed during the 365 days preceding the treatment index date. Medication covariates included medications for attention-deficit/hyperactivity disorder, analgesics, antiadrenergic medications, anticholinergics, antidiabetic medications, antihypertensives, antipsychotics, benzodiazepines, gastrointestinal medications, lipid-lowering medications, mood stabilizers, and sleep medications. Medication use was considered present if at least one qualifying dispensing record was observed during the baseline period. These demographic characteristics, comorbidities, and medication-use indicators were included as covariates in the propensity score models.

### 2.6 Statistical Analysis

For the primary analyses, six pairwise comparisons were evaluated among SSRIs, atypical antidepressants, SNRIs, and TCAs. A separate logistic propensity score model was estimated for each treatment pair using the prespecified baseline demographic, comorbidity, and medication covariates. Propensity score overlap weighting was used to estimate the average treatment effect in the overlap population (ATO) (Li et al., 2018). Covariate balance was assessed using SMDs, with absolute SMDs <0.10 considered indicative of negligible residual imbalance (Austin, 2009); effective sample sizes after weighting were also calculated. Associations with SREs were estimated using overlap-weighted Cox proportional hazards models with robust standard errors and Efron handling of tied event times and are reported as hazard ratios (HRs) with 95% confidence intervals (CIs). The proportional hazards assumption was evaluated using Schoenfeld residuals (Schoenfeld, 1982). Overlap-weighted Kaplan–Meier curves were used to describe SRE-free survival during as-treated follow-up. All tests were two-sided, with *P*<0.05 considered statistically significant.

Since the atypical antidepressant class included pharmacologically distinct agents, a secondary agent-specific analysis was conducted among bupropion, mirtazapine, and trazodone users. Individuals initiating more than one atypical antidepressant agent on the index date were excluded. Propensity scores and overlap weights were re-estimated for each agent-specific comparison using the same covariates and analytic approach as in the primary analysis.

Six sensitivity analyses evaluated alternative specifications of the new-user washout period, treatment-episode grace period, and maximum follow-up duration. The primary 180-day washout period was varied to 90 and 365 days, the 30-day grace period to 60 and 90 days, and the 2-year maximum follow-up to 1 and 3 years. Cohorts and treatment episodes were reconstructed under each specification, and the primary pairwise overlap-weighted analyses were repeated.

To assess variation in associations by age, analyses were stratified at age 18 years (<18 and ≥18 years), with propensity scores and overlap weights re-estimated within each age stratum for each pairwise comparison. Given the observed age-stratified pattern for atypical antidepressants versus SSRIs, further effect-modification analyses were conducted for this comparison using a treatment-by-age-group interaction (<18 vs ≥18 years), a linear treatment-by-continuous-age interaction, and a nonlinear spline specification for age.

Given evidence that clinical and behavioral features of ASD may differ by sex, we additionally evaluated sex as a potential effect modifier. For each primary treatment comparison, sex-stratified analyses were conducted among participants recorded as female or male, with propensity scores and overlap weights re-estimated separately within each sex. Formal effect modification was assessed using treatment-by-sex interaction terms in overlap-weighted Cox models.

## 3. Results

### 3.1 Study Population and Baseline Characteristics

The cohort selection and analytic workflow are summarized in **Figure 1**. The primary comparative analysis included 42,441 autistic individuals initiating one of four antidepressant classes, including 28,605 SSRI users, 9,756 atypical antidepressant users, 2,300 SNRI users, and 1,780 TCA users (shown in **Table 1**). Baseline characteristics differed across treatment groups before weighing. Overall, ages at treatment initiation ranged from 1.6 to 94 years. The mean age at treatment initiation varies from 20.9 years among SSRI users to 30.8 years among SNRI users. SNRI users had the highest prevalence of depression (39.6%) and anxiety (50.2%), while chronic pain and obesity were also more common in the SNRI and TCA groups. Baseline medication use differed across groups: ADHD medication and antipsychotic use were more common among atypical antidepressant users, whereas anticholinergic and gastrointestinal medication use were more frequent among TCA users.

**Figure 1.**
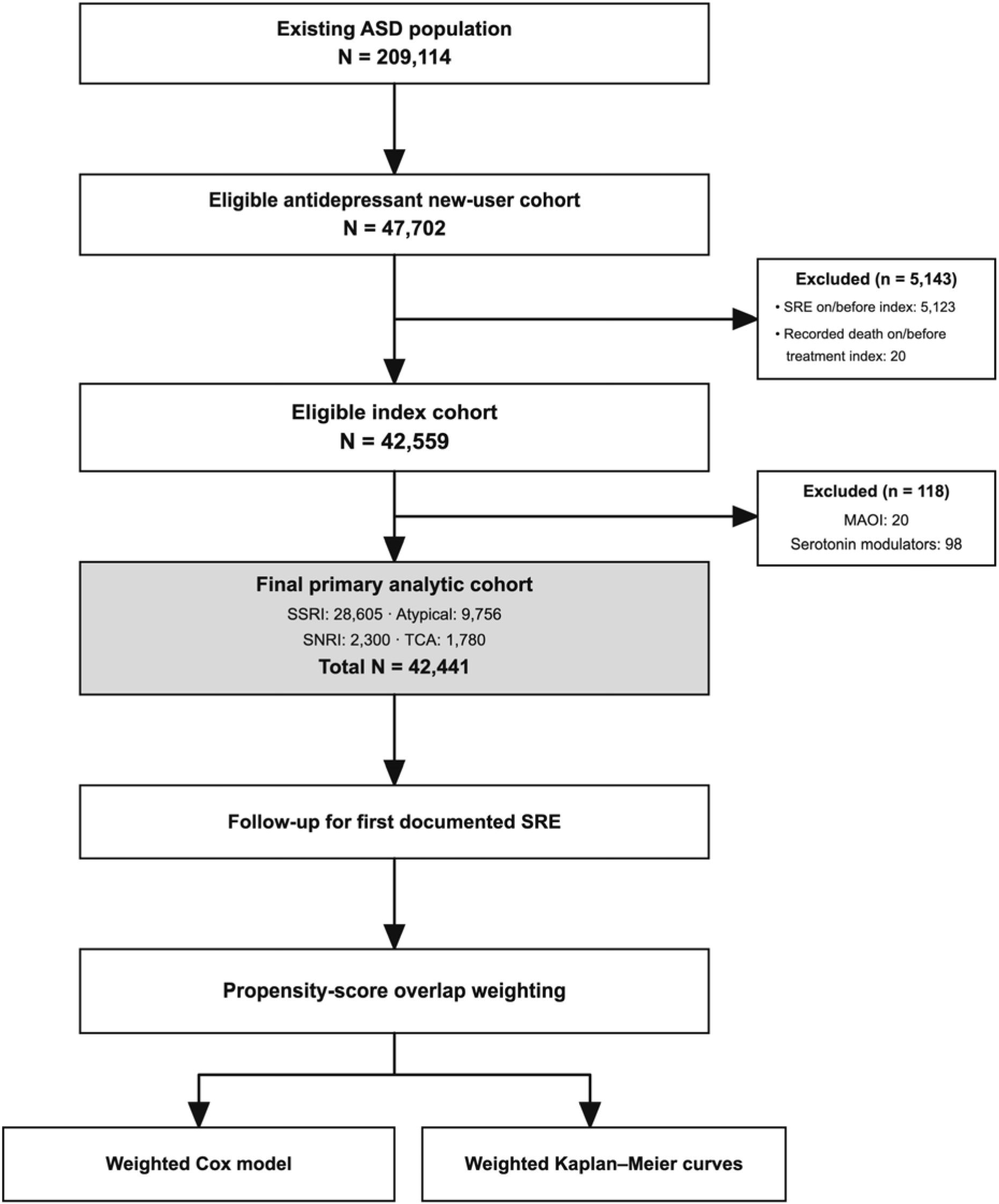
Cohort selection and analytic workflow for antidepressant new users with autism spectrum disorder

**Table 1.** Baseline Characteristics of Antidepressant Users With Autism Spectrum Disorder by Initial Treatment Class.

| **Characteristic** | **Atypical**<br>**(N=9,756)** | **SNRI**<br>**(N=2,300)** | **SSRI**<br>**(N=28,605)** | **TCA**<br>**(N=1,780)** |
| --- | --- | --- | --- | --- |
| **Age at index, mean (SD), years** | 24.52 (15.31) | 30.84 (15.68) | 20.91 (12.77) | 26.07<br>(14.83) |
| **Sex, Female (%)** | 2,958 (30.3) | 1,013 (44.0) | 9,092 (31.8) | 656 (36.9) |
| **Race, n (%)** |  |  |  |  |
| **Black or African American** | 1,081 (11.1) | 111 (4.8) | 1,943 (6.8) | 130 (7.3) |
| **White** | 7,314 (75.0) | 1,900 (82.6) | 22,454 (78.5) | 1,420 (79.8) |
| **Other** | 1,361 (13.9) | 289 (12.6) | 4,208 (14.7) | 230 (12.9) |
| **Co-occurring conditions, n (%)** |  |  |  |  |
| **Depression** | 1,842 (18.9) | 910 (39.6) | 5,494 (19.2) | 372 (20.9) |
| **ADHD** | 3,353 (34.4) | 752 (32.7) | 10,109 (35.3) | 524 (29.4) |
| **Substance use disorder** | 521 (5.3) | 182 (7.9) | 685 (2.4) | 95 (5.3) |
| **Schizophrenia spectrum disorders** | 478 (4.9) | 85 (3.7) | 608 (2.1) | 50 (2.8) |
| **Bipolar disorder** | 820 (8.4) | 186 (8.1) | 1,074 (3.8) | 138 (7.8) |
| **Anxiety** | 2,962 (30.4) | 1,155 (50.2) | 11,351 (39.7) | 636 (35.7) |
| **Obsessive-compulsive disorder** | 349 (3.6) | 145 (6.3) | 1,656 (5.8) | 110 (6.2) |
| **PTSD** | 370 (3.8) | 153 (6.7) | 773 (2.7) | 102 (5.7) |
| **Eating disorders** | 210 (2.2) | 39 (1.7) | 456 (1.6) | 43 (2.4) |
| **Personality disorders** | 328 (3.4) | 65 (2.8) | 656 (2.3) | 57 (3.2) |
| **Intellectual disability** | 946 (9.7) | 104 (4.5) | 1,901 (6.6) | 141 (7.9) |
| **Oppositional defiant or conduct disorder** | 755 (7.7) | 94 (4.1) | 2,096 (7.3) | 81 (4.6) |
| **Intermittent explosive disorder** | 120 (1.2) | 16 (0.7) | 268 (0.9) | 18 (1.0) |
| **Physical comorbidities, n (%)** |  |  |  |  |
| **Epilepsy** | 940 (9.6) | 138 (6.0) | 1,904 (6.7) | 182 (10.2) |
| **Chronic pain** | 717 (7.3) | 417 (18.1) | 1,275 (4.5) | 345 (19.4) |
| **Sleep-wake disorders** | 2,451 (25.1) | 506 (22.0) | 3,951 (13.8) | 404 (22.7) |
| **Obesity and overweight** | 1,343 (13.8) | 461 (20.0) | 3,362 (11.8) | 270 (15.2) |
| **Vision impairment** | 158 (1.6) | 43 (1.9) | 369 (1.3) | 41 (2.3) |
| **Hearing impairment** | 306 (3.1) | 70 (3.0) | 769 (2.7) | 75 (4.2) |
| **Sensory impairments** | 443 (4.5) | 107 (4.7) | 1,085 (3.8) | 108 (6.1) |
| **Baseline medications, n (%)** |  |  |  |  |
| **ADHD medication** | 3,778 (38.7) | 504 (21.9) | 9,766 (34.1) | 463 (26.0) |
| **Analgesic** | 1,496 (15.3) | 474 (20.6) | 3,216 (11.2) | 404 (22.7) |
| **Antiadrenergic** | 63 (0.6) | 17 (0.7) | 140 (0.5) | 15 (0.8) |
| **Anticholinergic** | 753 (7.7) | 157 (6.8) | 1,211 (4.2) | 293 (16.5) |
| **Antidiabetic** | 559 (5.7) | 201 (8.7) | 958 (3.3) | 95 (5.3) |
| **Antihypertensive** | 3,811 (39.1) | 647 (28.1) | 7,801 (27.3) | 550 (30.9) |
| **Antipsychotic** | 2,951 (30.2) | 455 (19.8) | 5,919 (20.7) | 379 (21.3) |
| **Benzodiazepine** | 130 (1.3) | 9 (0.4) | 220 (0.8) | 14 (0.8) |
| **GI medication** | 1,131 (11.6) | 374 (16.3) | 2,310 (8.1) | 355 (19.9) |
| **Lipid medication** | 606 (6.2) | 244 (10.6) | 1,039 (3.6) | 124 (7.0) |
| **Mood stabilizer** | 1,381 (14.2) | 263 (11.4) | 2,268 (7.9) | 220 (12.4) |
| **Sleep medication** | 349 (3.6) | 74 (3.2) | 444 (1.6) | 65 (3.7) |

*Values are presented as n (%) unless otherwise indicated. ADHD, attention-deficit/hyperactivity disorder; PTSD, post-traumatic stress disorder; SNRI, serotonin-norepinephrine reuptake inhibitor; SSRI, selective serotonin reuptake inhibitor; TCA, tricyclic antidepressant.*

After pairwise overlap weighting, covariate balance was substantially improved across all six comparisons, with maximum absolute standardized mean differences ranging from 0.002 to 0.075 (shown in **Supplementary Table 1**).

### 3.2 Primary Analysis

In the primary overlap-weighted Cox analyses, atypical antidepressant use was associated with a lower hazard of SRE compared with SSRI use (HR, 0.787; 95% CI, 0.632-0.979; *P*=0.032) (shown in **Figure 2**). TCA use was also associated with a lower hazard of SRE compared with SSRI use (HR, 0.487; 95% CI, 0.263-0.902; *P*=0.022).

**Figure 2.**
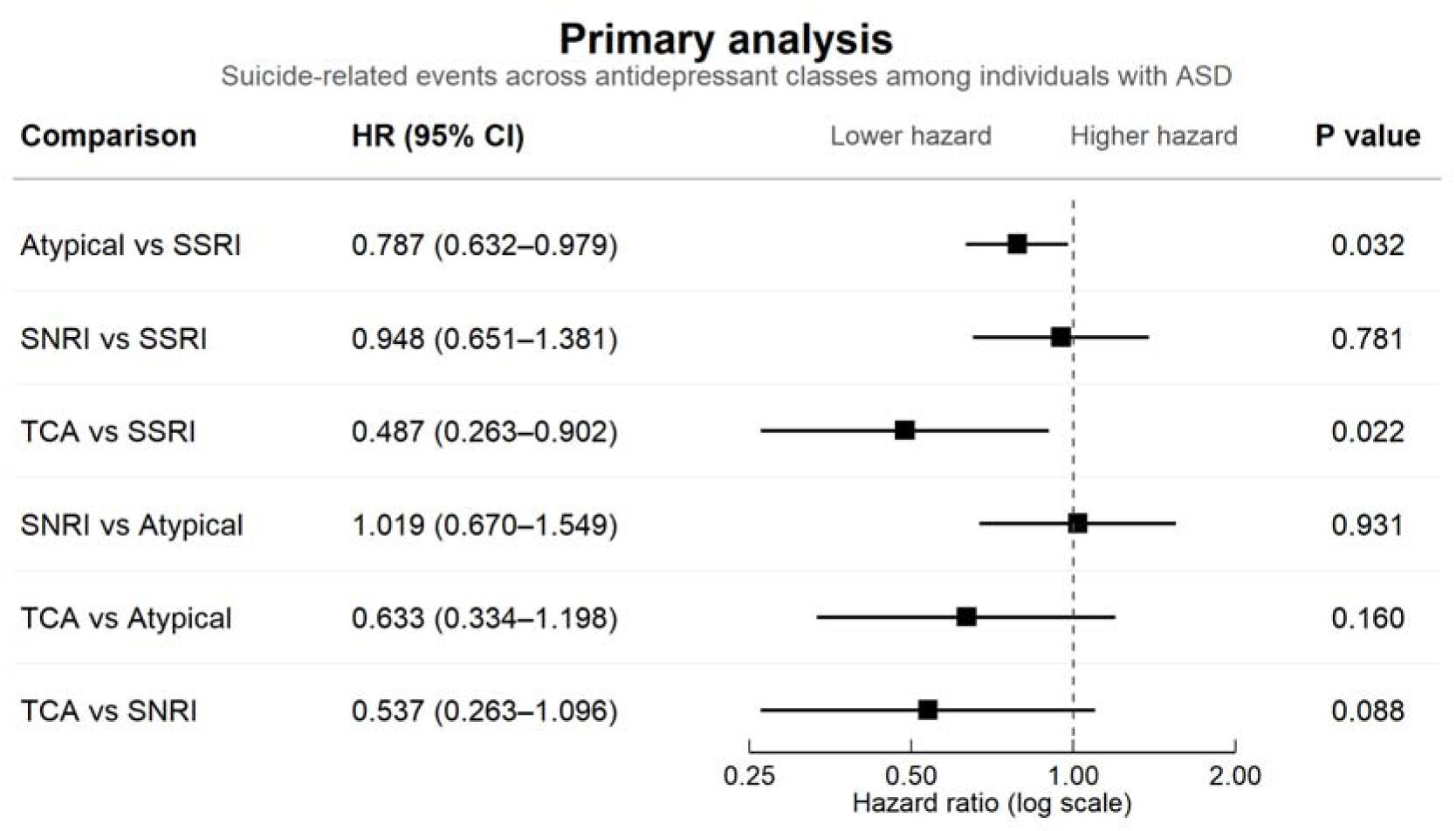
Pairwise Associations Between Antidepressant Treatment Classes and Suicide-Related Events

No statistically significant differences were observed for SNRI vs SSRI (HR, 0.948; 95% CI, 0.651-1.381), SNRI vs atypical antidepressants (HR, 1.019; 95% CI, 0.670-1.549), TCA vs atypical antidepressants (HR, 0.633; 95% CI, 0.334-1.198), or TCA vs SNRI (HR, 0.537; 95% CI, 0.263-1.096).

The weighted Kaplan-Meier curves for atypical antidepressants vs SSRIs and TCAs vs SSRIs are shown in **Figures** 3 **and 4**, respectively. Weighted Kaplan-Meier curves for the remaining pairwise comparisons are provided in the **Supplementary Figures 1-4**.

**Figure 3.**
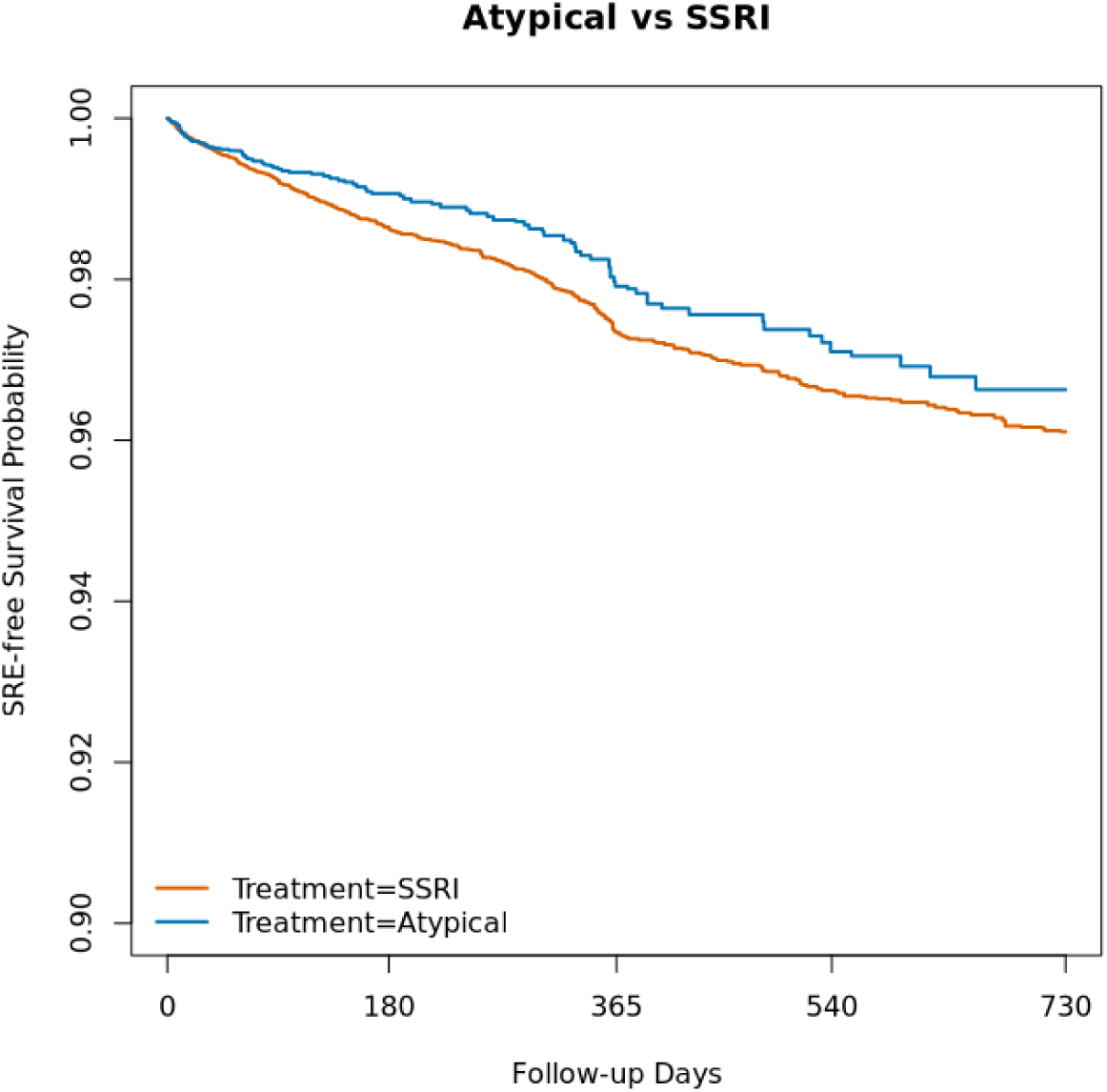
Overlap-Weighted Kaplan–Meier Curves for Suicide-Related Events: Atypical Antidepressants **vs.** SSRIs

**Figure 4.**
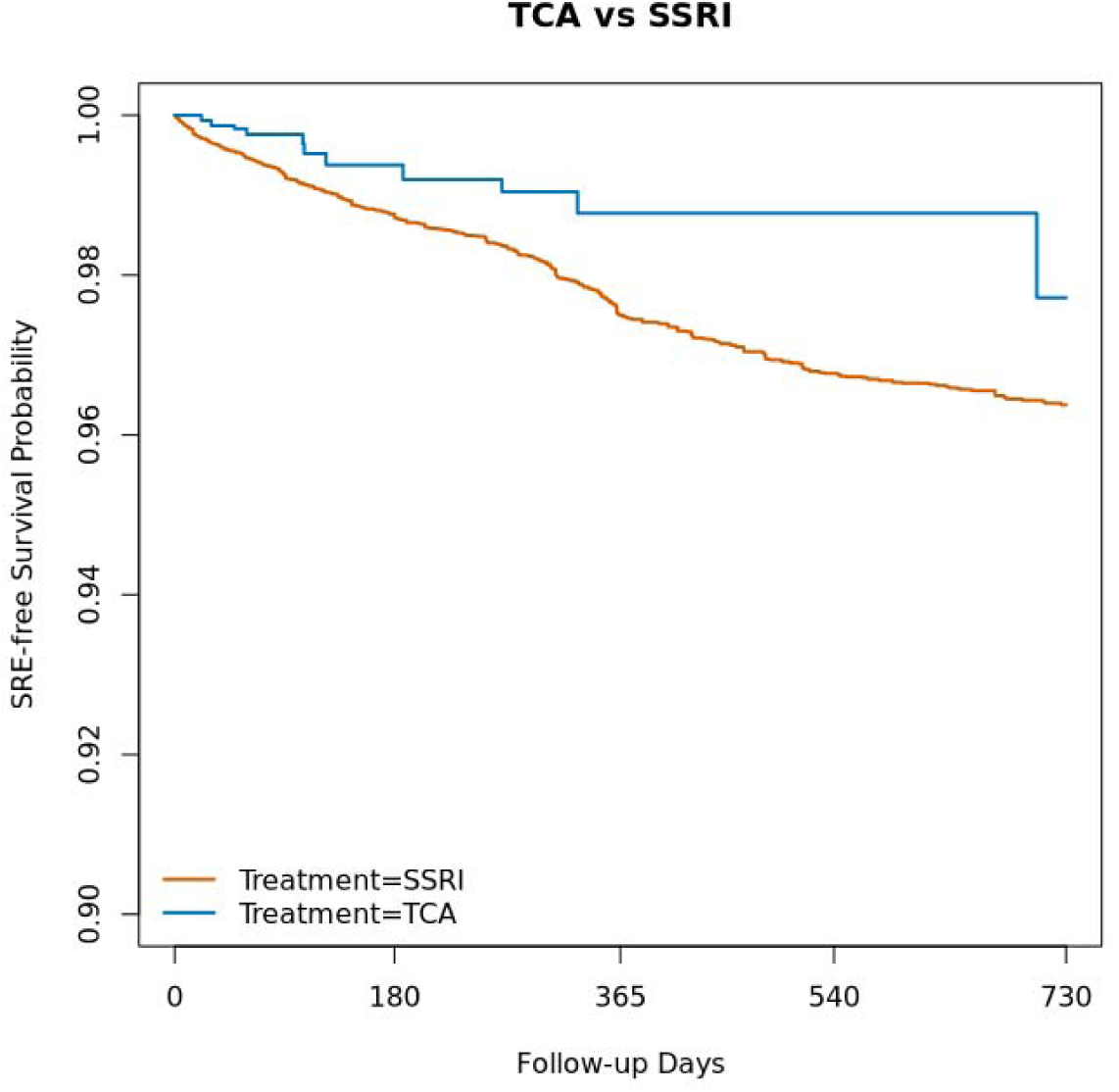
Overlap-Weighted Kaplan–Meier Curves for Suicide-Related Events: **TCAs vs.** SSRIs

### 3.3 Agent-Specific Analysis of Atypical Antidepressants

As the mechanism of action differs among specific atypical antidepressants, we performed a secondary analysis examining the association between individual atypical antidepressants and SSRI. Among the 9,756 individuals initiating atypical antidepressants, 5,858 (60.0%) initiated trazodone, 2,288 (23.5%) initiated bupropion, 1,601 (16.4%) initiated mirtazapine, and 9 (0.1%) initiated more than one atypical agent on the index date. The 9 individuals initiating multiple atypical antidepressants were excluded from the agent-specific analyses. Trazodone treatment was associated with a lower hazard of SRE compared with SSRI user (HR, 0.651; 95% CI, 0.484-0.877; *P*=0.005; shown in **Figure** 5). In contrast, no statistically significant differences relative to SSRIs were observed for bupropion (HR, 1.134; 95% CI, 0.792-1.624; *P*=0.491) or mirtazapine (HR, 0.854; 95% CI, 0.535-1.363; *P*=0.509). As shown in **Supplementary Table 2**, no statistically significant differences were observed in the post hoc comparisons of trazodone with SNRIs or TCAs.

**Figure 5.**
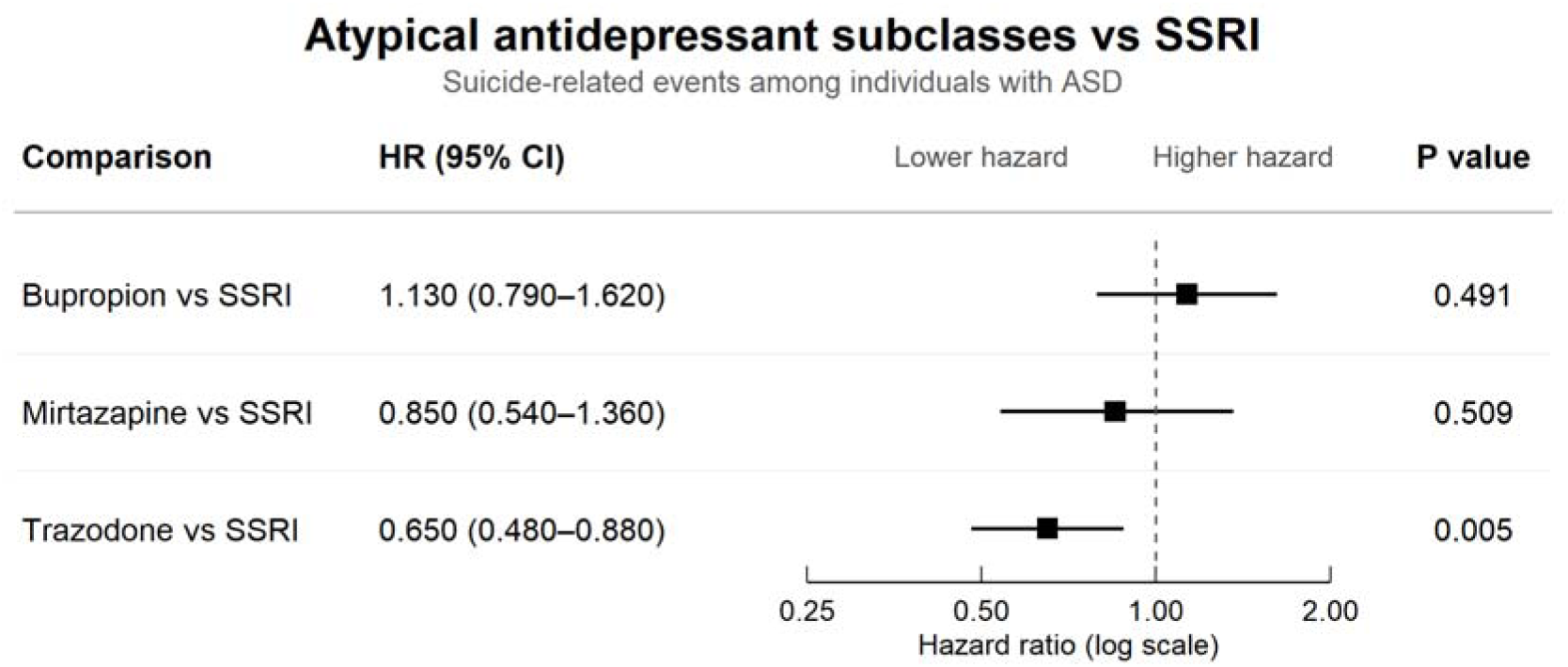
Agent-Specific Associations Between Atypical Antidepressants and Suicide-Related Events Compared with SSRIs

Weighted Kaplan–Meier curves for each of the three agent-specific comparisons are presented in the **Supplementary Figures 5-7**. No evidence of violation of the proportional hazard assumption was detected for these comparisons.

### 3.4 Sensitivity Analyses

Results were generally consistent across the six prespecified sensitivity analyses that changed the washout period, treatment episode grace period, and maximum follow-up duration (shown in **Supplementary Table** 3 and **Supplementary Figure 8**). For atypical antidepressants vs SSRIs, hazard ratio estimates remained below 1.0 across all sensitivity analyses, ranging from 0.700 to 0.838. In the sensitivity analysis using a 90-day new-user washout period (Sensitivity Analysis 1), the association did not reach statistical significance (HR, 0.838; 95% CI, 0.684–1.027; *P* = 0.089), whereas the association remained statistically significant in the other five sensitivity analyses. For TCAs versus SSRIs, the association remained statistically significant across all six sensitivity analyses, with hazard ratio estimates ranging from 0.468 to 0.549. No statistically significant associations were observed for the other four pairwise comparisons across the sensitivity analyses

### 3.5 Age- and Sex-Stratified and Effect-Modification Analyses

To evaluate whether associations between antidepressant class and SRE risk differed by age, we examined age 18 years as a clinically relevant threshold distinguishing pediatric and adult populations. In analyses stratified at 18 years old, atypical antidepressant use was associated with a lower hazard of SRE compared with SSRI user among individuals younger than 18 years (HR, 0.679; 95% CI, 0.488-0.944; *P*=0.021), whereas the corresponding estimate among individuals aged 18 years or older was 0.963 (95% CI, 0.712-1.303; P=0.806) (shown in **Table 2**). None of the other five pairwise comparisons showed statistically significant associations within either age stratum.

**Table 2.** Age-Stratified Pairwise Associations Between Antidepressant Treatment Classes and Suicide-Related Events.

| Comparison pairs | <18 years old |  | ≥18 years old |  |
| --- | --- | --- | --- | --- |
|  | HR (95% CI) | P value | HR (95% CI) | P value |
| **Atypical vs SSRI** | 0.679 (0.488–0.944) | 0.021 | 0.963 (0.712–1.303) | 0.806 |
| **SNRI vs SSRI** | 1.180 (0.654–2.131) | 0.582 | 0.849 (0.521–1.384) | 0.512 |
| **TCA vs SSRI** | 0.485 (0.177–1.328) | 0.159 | 0.529 (0.242–1.156) | 0.111 |
| **SNRI vs Atypical** | 1.543 (0.784–3.038) | 0.209 | 0.824 (0.483–1.404) | 0.476 |
| **TCA vs Atypical** | 0.773 (0.272–2.199) | 0.630 | 0.577 (0.260–1.284) | 0.178 |
| **TCA vs SNRI** | 0.451 (0.142–1.428) | 0.176 | 0.654 (0.265–1.612) | 0.356 |

Given the observed difference in the age-stratified estimates for atypical antidepressants vs SSRIs, further effect-modification analyses were conducted for this comparison. When age was dichotomized as <18 versus ≥18 years, the interaction between atypical antidepressant versus SSRI treatment and age was statistically significant (*P* for interaction=0.037). However, the interaction between atypical antidepressant versus SSRI treatment and age was not statistically significant when age was modeled as a continuous linear variable (*P* for interaction=0.113) or using a nonlinear spline (*P*=0.267) (shown in **Supplementary Table** 4).

In sex-stratified analyses, atypical antidepressants were associated with a lower observed hazard of SREs relative to SSRIs among female participants (HR, 0.60; 95% CI, 0.39 - 0.91), whereas the corresponding estimate among male participants was closer to the null (HR, 0.88; 95% CI, 0.68 - 1.14). However, the treatment-by-sex interaction was not statistically significant (P for interaction = 0.098). No clear evidence of treatment-by-sex interaction was observed for the other primary pairwise comparisons (all P for interaction > 0.05; shown in **Supplementary Table 5**).

## 4. Discussion

This study provides new real-world evidence on the comparative risk of suicide-related events (SREs) across antidepressant treatments among 42,441 autistic people utilizing antidepressants in the Truveta database. In pairwise propensity score overlap-weighted analyses with as-treated follow-up, TCA and atypical antidepressant treatment were associated with a lower observed hazard of SREs compared with SSRI treatment, whereas no statistically significant difference was observed between SNRI and SSRI treatment. Similar estimates for TCAs were observed across sensitivity analyses with different washout period, grace period, and maximum follow-up duration. However, the lower hazard observed for atypical antidepressants as a class was not consistent across the individual agents included in this category. In agent-specific analyses, trazodone treatment was associated with a lower observed hazard of SREs compared with SSRI treatment, whereas no statistically significant differences were observed for bupropion or mirtazapine.

Comparative evidence on suicide-related outcomes across antidepressant treatments in autistic populations remains limited. The present study broadens the existing evidence in autism research by using a new-user, active-comparator design to examine head-to-head associations between commonly used antidepressant treatments and a broader range of SREs under as-treated follow-up. In the absence of directly comparable evidence from autistic populations, the interpretation of our findings therefore draws primarily on studies conducted among individuals with depressive disorders. TCA treatment was associated with a lower observed SRE hazard compared with SSRI treatment (HR, 0.487; 95% CI, 0.263–0.902), and the direction of this association was similar across the prespecified sensitivity analyses. This finding is consistent with the lower risk reported by Su et al. (2019) in individuals with depressive disorders but differs from the little or no difference reported in other depression cohorts (Coupland et al., 2015; Schneeweiss et al., 2010). TCAs are also prescribed for nonpsychiatric indications, such as chronic pain and migraine (Wong et al., 2017), and their adverse-effect profile and toxicity in overdose may influence treatment selection (Taylor et al., 2024). In our cohort, chronic pain and sleep-wake disorders were more common among TCA users than SSRI users before weighting, suggesting potentially different prescribing contexts. Although these measured characteristics were balanced after weighting, unmeasured differences in treatment indication and symptom severity may have contributed to the observed association and may also complicate comparisons with depression cohorts.

We observed no statistically significant difference in SRE hazard between SNRI and SSRI treatment (HR, 0.948; 95% CI, 0.651–1.381). Among previous studies conducted in depressive-disorder populations, Miller et al. (2014) offers the closest methodological parallel, as it similarly used a new-user, active-comparator design with as-treated follow-up, although it assessed deliberate self-harm as the outcome. Its finding of similar risk between SNRIs and SSRIs was consistent with those of Schneeweiss et al. (2010) and Valuck et al. (2016), whereas Su et al. (2019) and Wu et al. (2017) reported higher risks among SNRI users. In addition, prior authors have noted that SNRIs may be preferentially prescribed to patients with more severe, chronic, or treatment-resistant depression, leaving the possibility of residual confounding by treatment selection (Valuck et al., 2016). Accordingly, our finding adds ASD-specific evidence but does not establish equivalence between SNRIs and SSRIs.

Atypical antidepressant treatment was associated with a lower observed SRE hazard than SSRI treatment at the class level (HR, 0.787; 95% CI, 0.632–0.979). However, our atypical antidepressant category comprised three pharmacologically distinct agents: bupropion, mirtazapine, and trazodone, which supported a further separate evaluation. In agent-specific analyses, a lower observed SRE hazard relative to SSRIs was observed for trazodone (HR, 0.651; 95% CI, 0.484–0.877), whereas no clear differences were observed for bupropion (HR, 1.134; 95% CI, 0.792–1.624) or mirtazapine (HR, 0.854; 95% CI, 0.535–1.363). Additional post hoc comparisons did not provide clear evidence of differences between trazodone and either SNRIs or TCAs.

Previous studies have likewise not consistently treated bupropion, mirtazapine, and trazodone as a single antidepressant class, and agent-specific findings have varied across depression cohorts. For mirtazapine, Wu et al. (2017) and Coupland et al. (2015) reported higher risks of self-harm outcomes relative to fluoxetine and citalopram, respectively. However, in an active-comparator study of second-line antidepressant treatment, the initially higher rate of serious self-harm among mirtazapine users was attenuated after adjustment for baseline differences, with no statistically significant difference relative to SSRIs (Joseph et al., 2022). This attenuation suggests that differences in baseline clinical risk and the selection of mirtazapine as a later-line treatment may partly explain the higher estimates reported in some earlier studies. For bupropion, Wu et al. (2017) reported a lower hazard of self-harm hospitalization. However, evidence for bupropion remains limited because many previous observational studies did not include it, possibly owing to the difficulty of distinguishing its use for depression from its use for smoking cessation (Wu et al., 2017). Schneeweiss et al. (2010), for example, explicitly excluded bupropion for this reason. Evidence for trazodone has been similarly inconsistent. Wu et al. (2017) observed no statistically significant difference in self-harm hospitalization relative to fluoxetine, Coupland et al. (2015) reported a higher hazard of attempted suicide or self-harm relative to citalopram, and Olfson et al. (2006) reported a lower but statistically nonsignificant estimate relative to no antidepressant treatment. Together, these findings do not indicate a consistent trazodone-associated risk across populations and treatment comparators. Trazodone antagonizes 5-HT2A receptors and inhibits serotonin reuptake in a dose-dependent manner (Fagiolini et al., 2023), although whether these actions explain the lower observed SRE hazard remains unclear. Trazodone is also frequently prescribed at low doses for insomnia, including outside the treatment of depression (Fagiolini et al., 2023; Wong et al., 2017). Differences in prescribing indications may reflect underlying differences in psychiatric risk between trazodone and SSRI users, potentially contributing to the observed association. Further studies incorporating treatment indication, dose, and psychiatric symptom severity are needed to distinguish among these explanations.

Our age-stratified analysis suggested that the lower SRE hazard associated with atypical antidepressants relative to SSRIs was more apparent among individuals younger than 18 years. Age-dependent associations with suicidality have also been reported in the broader antidepressant literature (Stone et al., 2009). Although the binary age-group interaction was statistically significant, interaction analyses treating age as a continuous or nonlinear variable did not provide consistent evidence of effect modification. The age-stratified findings should therefore be considered exploratory. Although sex has been identified as a potentially important source of heterogeneity in autism, we found no clear evidence that the associations between antidepressant treatment class and SREs differed by sex.

This study has several strengths. The large, geographically diverse Truveta cohort enabled head-to-head evaluation of commonly used antidepressant treatments in an ASD population for which comparative evidence remains limited. The new-user, active-comparator design and pairwise propensity score overlap weighting improved comparability between treatment groups across a broad range of measured baseline characteristics. As-treated follow-up censored individuals at treatment discontinuation or initiation of a different antidepressant class according to prespecified rules, while sensitivity analyses varying key exposure and follow-up definitions assessed the robustness of the findings. Agent-specific analyses further allowed us to examine whether the class-level association for atypical antidepressants was consistent across its pharmacologically distinct components.

Several limitations should be considered. Despite the active-comparator design and balance in measured baseline characteristics, residual confounding by treatment indication, psychiatric symptom severity, and other factors influencing treatment selection remains possible. In particular, the EHR data did not provide dimensional measures of autism-related characteristics, cognitive ability, or speaking ability. These factors may influence both treatment selection and the recognition or documentation of suicide-related symptoms and events, and therefore represent additional potential sources of residual confounding and outcome misclassification. Medication dispensing indicates that treatment was available but does not establish adherence. Treatment exposure and discontinuation were therefore operationally defined using recorded days’ supply and the prespecified grace period rather than directly observed medication use, potentially resulting in exposure misclassification. Moreover, censoring at discontinuation or treatment switching could have been informative if these changes were related to emerging symptoms or suicide risk. SRE ascertainment relied on diagnoses recorded within participating health systems, and events occurring elsewhere or not resulting in clinical attention may have been missed. We did not separately evaluate associations with individual SRE components, which may differ in their relationships with antidepressant treatment. The relatively small TCA Sensitivity analyses’ consistent results should therefore be viewed as consistency across alternative analytic specifications rather than independent confirmation of the association. Agent-specific and post hoc findings should likewise be interpreted cautiously because they were based on fewer events and multiple comparisons. Finally, pairwise overlap-weighted estimates pertain to the populations represented in the respective treatment comparisons and should not be directly compared across treatment pairs. The findings should therefore be interpreted as observational associations rather than causal effects.

## 5. Conclusion

Among autistic people in this database, TCA treatment was associated with a lower observed SRE hazard than SSRI treatment during as-treated follow-up, and this finding remained consistent across sensitivity analyses. A lower observed hazard was also found for atypical antidepressants as a class; however, agent-specific analyses showed this association for trazodone but not for bupropion or mirtazapine. No statistically significant difference was observed between SNRI and SSRI treatment. These findings provide autism-specific observational evidence regarding comparative SRE hazards across commonly used antidepressant treatments.

## Data Availability

The data that support the findings of this study were obtained from Truveta and are subject to third-party restrictions. The authors are not permitted to share the underlying patient-level data. Access to Truveta data may be obtained directly from Truveta to its applicable data access requirements and agreements.

https://www.truveta.com/

## Acknowledgments

Research reported in this publication was supported by the National Institute Of Mental Health under Award Number P50MH130957. The content is solely the responsibility of the authors and does not necessarily represent the official views of the National Institutes of Health.

