## Supplement Table 1 for "Comparative Risk of Suicide-Related Events Across Antidepressant Classes in Autistic Individuals: A New-User Active Comparator Cohort Study"

**Supplementary table 1** Covariate Balance After Pairwise Propensity Score Overlap Weighting

| Comparison pair | Max SMD |
| --- | --- |
| Atypical vs SSRI | 0.019 |
| SNRI vs SSRI | 0.075 |
| TCA vs SSRI | 0.057 |
| SNRI vs Atypical | 0.044 |
| TCA vs Atypical | 0.004 |
| TCA vs SNRI | 0.002 |

*SMDs represent standardized mean differences across antidepressant treatment groups after overlap weighting.*

**Supplementary figure 1** Overlap-Weighted Kaplan–Meier Curves for Suicide-Related Events: **SNRIs vs.** SSRIs

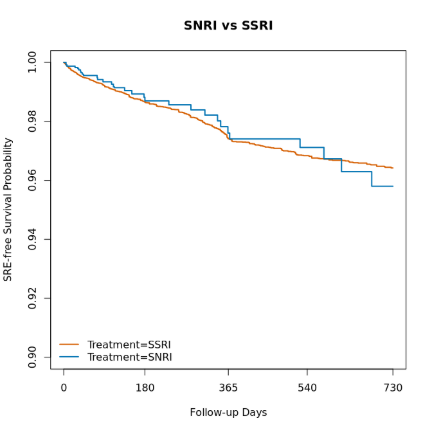

**Supplementary figure 2** Overlap-Weighted Kaplan–Meier Curves for Suicide-Related Events: **SNRIs vs. Atypical Antidepressant**

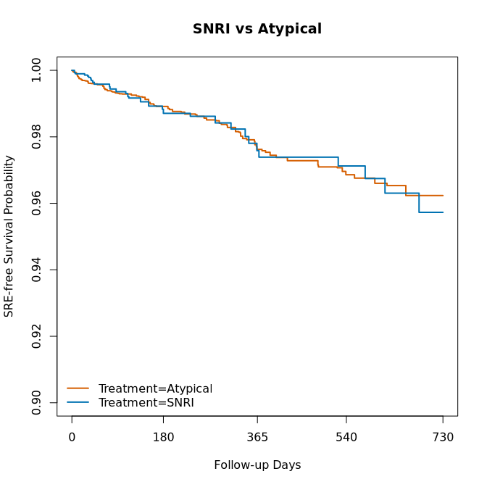

**Supplementary figure 3** Overlap-Weighted Kaplan–Meier Curves for Suicide-Related Events: **TCAs vs. Atypical Antidepressant**

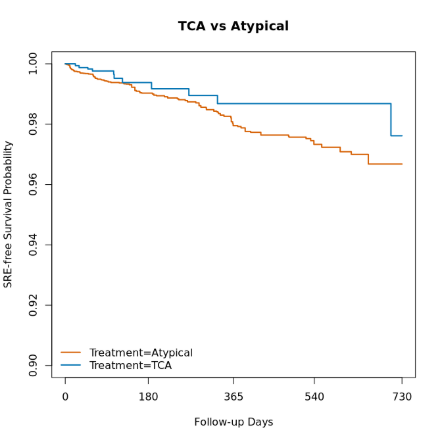

**Supplementary figure 4** Overlap-Weighted Kaplan–Meier Curves for Suicide-Related Events: **TCAs vs. SNRIs**

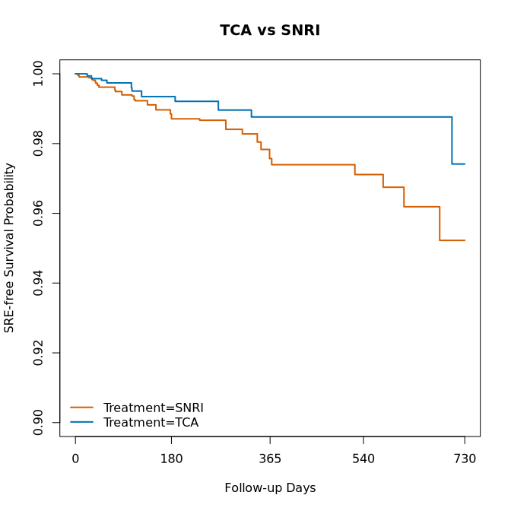

**Supplementary table 2** Post Hoc Comparisons of Trazodone With SNRIs and TCAs for Suicide-Related Events

| **Comparison** | **HR (95% CI)** | **P value** |
| --- | --- | --- |
| Trazodone vs SNRI | 0.927 (0.562–1.528) | 0.766 |
| Trazodone vs TCA | 1.235 (0.627–2.434) | 0.541 |

**Supplementary figure 5** Overlap-Weighted Kaplan–Meier Curves for Suicide-Related Events: **Bupropion** **vs.** SSRIs

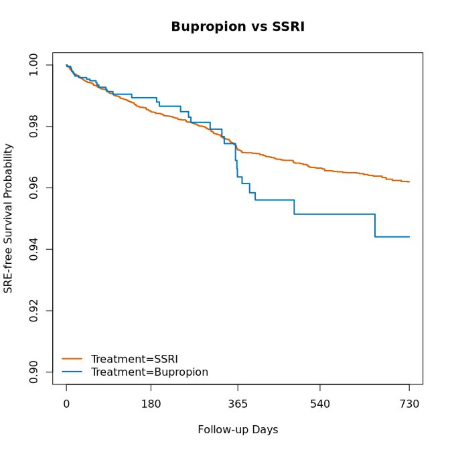

**Supplementary figure 6** Overlap-Weighted Kaplan–Meier Curves for Suicide-Related Events: **Mirtazapine vs.** SSRIs

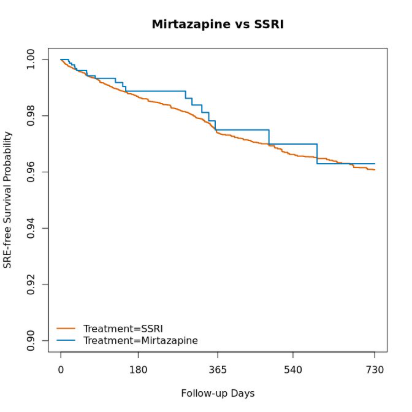

**Supplementary figure 7** Overlap-Weighted Kaplan–Meier Curves for Suicide-Related Events: **Trazodone vs.** SSRIs

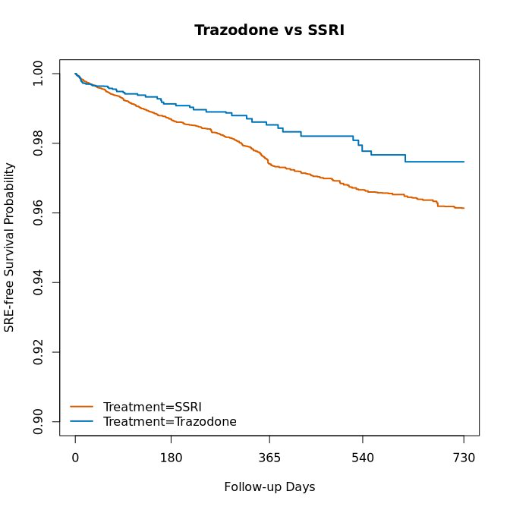

**Supplementary table3** Alternative Analysis Specifications Used in Sensitivity Analyses

| Scenario | Washout (days) | Grace (days) | Follow-up (days) |
| --- | --- | --- | --- |
| Primary analysis | 180 | 30 | 730 |
| Sensitivity analysis 1 | 90 | 30 | 730 |
| Sensitivity analysis 2 | 365 | 30 | 730 |
| Sensitivity analysis 3 | 180 | 60 | 730 |
| Sensitivity analysis 4 | 180 | 90 | 730 |
| Sensitivity analysis 5 | 180 | 30 | 1095 |
| Sensitivity analysis 6 | 180 | 30 | 365 |

**Supplementary figure 8** Sensitivity Analyses of Pairwise Associations Between Antidepressant Treatment Classes and Suicide-Related Events

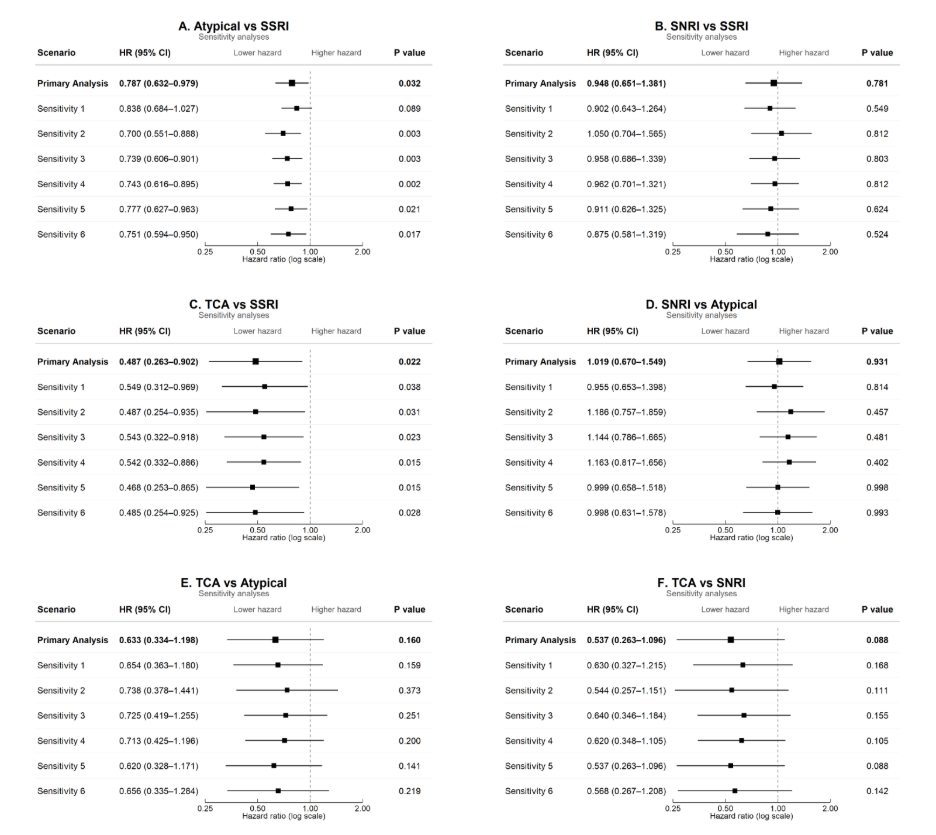

**Supplementary table 4** Age Interaction Analyses for Atypical Antidepressants **vs.** SSRIs

| **Age modeling approach** | **Result** |
| --- | --- |
| Binary age interaction: < 18 vs ≥ 18 | P-interaction = 0.037 |
| Continuous linear age interaction | P-interaction = 0.113 |
| Continuous spline age interaction | P-interaction = 0.267 |

**Supplementary table 5** Sex-stratified associations between antidepressant treatment class and suicide-related events

| **Comparison** | **Female HR (95% CI)** | **Male HR (95% CI)** | **P for interaction** |
| --- | --- | --- | --- |
| Atypical vs SSRI | 0.60 (0.39–0.91) | 0.88 (0.68–1.14) | 0.098 |
| SNRI vs SSRI | 0.92 (0.53–1.58) | 0.96 (0.57–1.62) | 0.738 |
| TCA vs SSRI | 0.59 (0.23–1.51) | 0.43 (0.19–0.98) | 0.736 |
| SNRI vs Atypical | 1.31 (0.68–2.53) | 0.89 (0.51–1.56) | 0.483 |
| TCA vs Atypical | 0.92 (0.33–2.57) | 0.49 (0.21–1.15) | 0.362 |
| TCA vs SNRI | 0.70 (0.24–2.04) | 0.41 (0.15–1.09) | 0.495 |
